# Multi-omics characterization of type 2 diabetes associated genetic variation

**DOI:** 10.1101/2024.07.15.24310282

**Authors:** Ravi Mandla, Kim Lorenz, Xianyong Yin, Ozvan Bocher, Alicia Huerta-Chagoya, Ana Luiza Arruda, Anthony Piron, Susanne Horn, Ken Suzuki, Konstantinos Hatzikotoulas, Lorraine Southam, Henry Taylor, Kaiyuan Yang, Karin Hrovatin, Yue Tong, Maria Lytrivi, Nigel W. Rayner, James B. Meigs, Mark I. McCarthy, Anubha Mahajan, Miriam S. Udler, Cassandra N. Spracklen, Michael Boehnke, Marijana Vujkovic, Jerome I. Rotter, Decio L. Eizirik, Miriam Cnop, Heiko Lickert, Andrew P. Morris, Eleftheria Zeggini, Benjamin F. Voight, Josep M. Mercader

## Abstract

Discerning the mechanisms driving type 2 diabetes (T2D) pathophysiology from genome-wide association studies (GWAS) remains a challenge. To this end, we integrated omics information from 16 multi-tissue and multi-ancestry expression, protein, and metabolite quantitative trait loci (QTL) studies and 46 multi-ancestry GWAS for T2D-related traits with the largest, most ancestrally diverse T2D GWAS to date.

Of the 1,289 T2D GWAS index variants, 716 (56%) demonstrated strong evidence of colocalization with a molecular or T2D-related trait, implicating 657 *cis*-effector genes, 1,691 distal-effector genes, 731 metabolites, and 43 T2D-related traits. We identified 773 of these *cis-* and distal-effector genes using either expression QTL data from understudied ancestry groups or inclusion of T2D index variants enriched in underrepresented populations, emphasizing the value of increasing population diversity in functional mapping. Linking these variants, genes, metabolites, and traits into a network, we elucidated mechanisms through which T2D-associated variation may impact disease risk. Finally, we showed that drugs targeting effector proteins were enriched in those approved to treat T2D, highlighting the potential of these results to prioritize drug targets for T2D.

These results represent a leap in the molecular characterization of T2D-associated genetic variation and will aid in translating genetic findings into novel therapeutic strategies.

## Introduction

Type 2 diabetes (T2D) and its associated complications are one of the biggest global health problems of the 21^st^ century^1,2^. The biological mechanisms underlying T2D are not fully understood, yet expanding upon pathways for T2D could inform therapeutic approaches. It has been demonstrated that clinical trials of drugs that have genetic support are more likely to be successful and to be granted expedited development and review by the FDA^3–6^. Therefore, a logical approach to accelerate development of new T2D drugs is through linking T2D-associated variants with their effector genes.

One strategy to nominate potential disease effector genes is to identify rare variants that alter the protein coding sequence through putative loss-of-function (pLOF) or amino-acid altering variants^7,8^. While this approach has generated numerous leads^9^, discovery of rare coding variants associated with T2D has been limited by small sample sizes from available exome or genome sequencing studies, especially in underrepresented populations. In contrast, large-scale multi-ancestry array-based genome-wide association studies (GWAS) have been successful in identifying hundreds of genetic associations^10–15^. However, only 50 (4%) of the 1,289 T2D-associated genetic variants from the latest T2D GWAS are themselves a pLOF or amino-acid altering variant^15^. Previous results have also suggested only 43% of T2D-associated protein-altering variants show evidence of causality after fine-mapping^16^, complicating efforts with this strategy alone to identify the effector genes underlying most T2D associations. Additionally, inference of the tissue of action, effect direction, and physiological mechanisms of T2D-associated variants are also important to translate these discoveries into mechanistic insights and novel drug targets.

Many T2D variant-to-function efforts have attempted to determine these downstream targets through the identification of shared causal signals underlying both T2D and a single omics layer from a quantitative trait loci (QTL) analysis, with gene expression being the most common^17–20^. However, a recent GWAS for T2D could only map *cis*-effector genes for 21% of their signals when using transcriptomic and proteomic data^11^, motivating the pursuit of integrating larger and more ancestrally diverse molecular data to identify additional mechanisms. To date, few studies in T2D have attempted to jointly analyze transcriptomic, proteomic, and metabolomic data to link genetic variations to T2D biological pathways. Furthermore, there have been limited attempts to explore the overlap of these functional genomics datasets with cardiometabolic trait-associated clusters for T2D^15,21^ to better characterize the biology underlying T2D heterogeneity. More importantly, previous analyses have focused on populations genetically similar to European ancestry individuals from the 1000 Genomes Project (1000G), which we henceforth refer to as EUR-like, limiting our understanding of the biological downstream effects of T2D in other population groups such as those genetically similar to African ancestry (AFR-like), admixed-American (AMR-like), East Asian ancestry (EAS-like), and South asian ancestry (SAS-like).

Here, we tested for colocalization between the largest, recently reported multi-ancestry T2D GWAS meta-analysis^15^ with association datasets for transcriptomic, proteomic, and metabolomic traits to identify effector transcripts and downstream targets for T2D across multiple tissues and ancestry groups. Additionally, we used GWAS data from T2D-related traits, including glycemic, cardiometabolic, and anthropometric traits, to understand the physiological mechanisms linked to T2D-associated variation and to further characterize the heterogeneity of T2D loci. We found evidence of colocalization between T2D signals and molecular or T2D-related traits for the majority of the T2D signals, expanding the catalog of variant-to-gene mappings and physiologic correlates with disease. We combined these associations into an interactive network to define relationships between genetic variants, molecular trait data, and T2D-related traits, providing a resource to prioritize mechanisms and propose potential therapeutic hypotheses.

## Results

### Overall analysis strategy

To understand molecular mechanisms and physiological mechanisms of T2D-associated genetic variation, we evaluated evidence of pairwise-colocalization between each of the 1,289 T2D-associated index variants identified in Suzuki et al.^15^ and *cis-*gene expression QTL from diverse tissues and ancestries (eQTL, 10 datasets), protein QTL (*cis-* and *trans-*pQTL, 4 datasets), metabolite QTL (metabQTL, 2 datasets), and T2D-related cardiometabolic trait GWAS (46 datasets; **Fig. 1a**; **Supplementary Tables 1** and **2**; **Methods**). We identified 12,180 colocalizations with a posterior probability of a shared causal variant (PP.H4) > 0.8, corresponding to a total of 716 T2D index variants colocalizing with 657 *cis*-effector genes from an eQTL or *cis*-pQTL, 1,691 distal-effector genes from *trans*-pQTL, 731 metabolites, and 43 T2D-related traits (**Fig. 1b**, **1c**; **Supplementary Tables 3**, **4**, **5**, and **6**; **Methods**).

**Figure 1:**
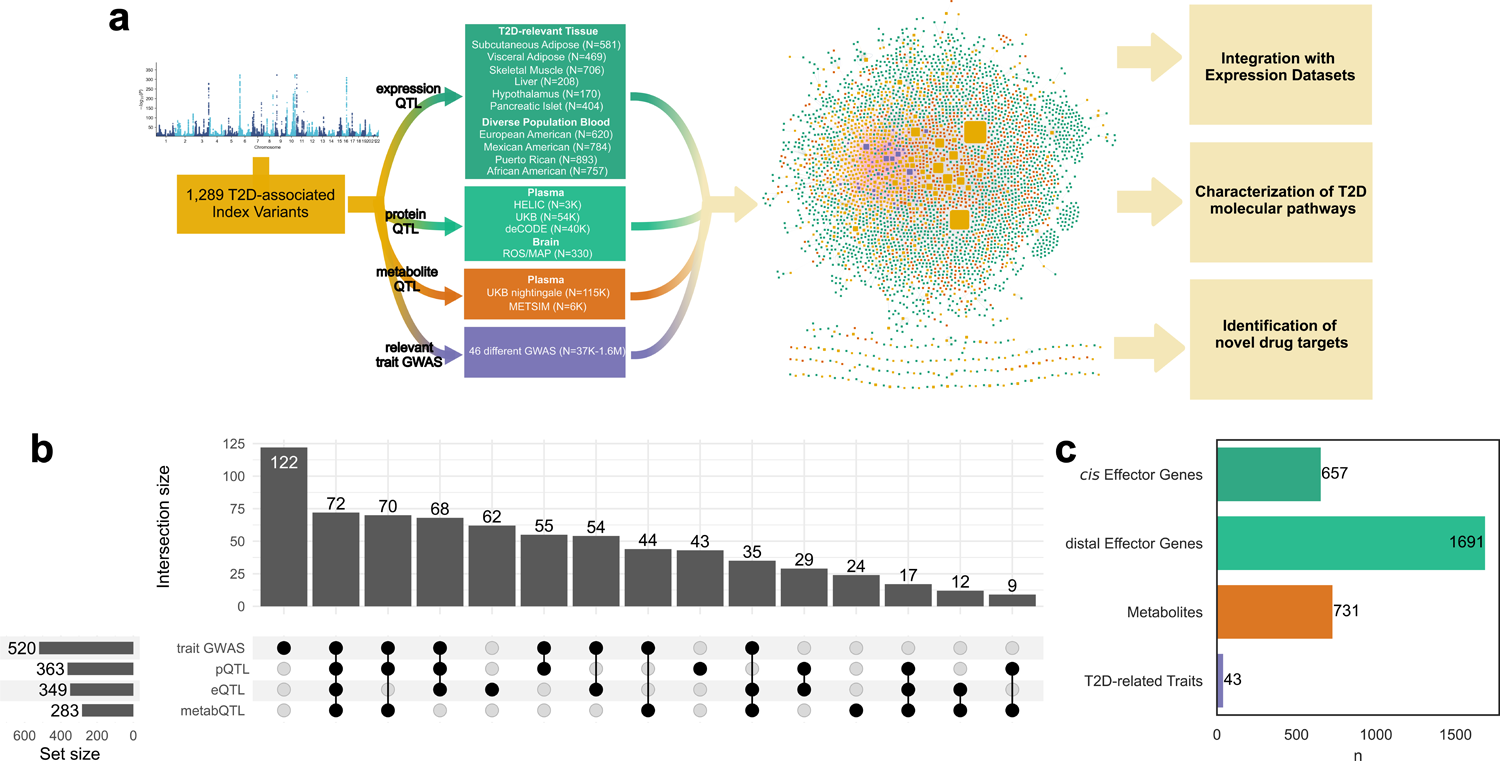
Overview of Project. **a)** Genomic regions containing the 1,289 T2D-associated index variants from Suzuki et al.^15^ were tested for evidence of colocalization with 10 eQTL datasets, 4 pQTL datasets, 2 metabQTL datasets, and 46 related trait GWAS. Colocalizations were then mapped to an interactive network to visualize genes (from an eQTL or pQTL, colored green), metabolites (colored orange), or traits (colored purple) with evidence of sharing a causal variant with T2D around a index variant (colored yellow). These results were then used in downstream analyses to identify enrichment with expression datasets, better understand T2D pathways, and identify drug targets for T2D. **b)** Upset plot of the 716 T2D index variants mapped to an association in an eQTL, pQTL, metabQTL, or trait GWAS dataset (with a colocalization PP.H4 >0.8, Methods). **c)** Bar plot of the number of effector genes, metabolites, and traits identified from colocalization analyses with T2D.

We represented the 12,180 colocalizations as a multi-layer colocalization network, where nodes represent T2D GWAS index variants, genes, metabolites, or traits, and edges represent a colocalization between T2D and eQTLs, pQTLs, metabQTLs or trait GWAS data (**Fig. 1a**; **Supplementary File**).

### Identification of T2D cis-effector genes and their relevant tissues

To identify candidate T2D effector genes and their tissues of action, we tested for evidence of colocalization between T2D and eQTL datasets from blood^22,23^ and six T2D-related tissues: pancreatic islets^19^, subcutaneous and visceral adipose tissue, liver, hypothalamus, and skeletal muscle^23^ (**Methods**). We observed 1,324 colocalizations, representing 632 candidate *cis*-effector genes, and 349 (27.1%) of the 1,289 T2D index variants. Of the 349 index variants, 110 (32.0%) colocalized to different effector genes in different tissues, suggesting evidence of varying effects by tissue (**Supplementary Table 7**).

To quantify the improvement in our list of T2D effector genes, we compared our results to previous studies that performed colocalization analyses on different T2D GWAS^11,14,19,24^. We found that the present analysis doubles the number of candidate effector genes compared to the largest previous study^14^, owing to the larger sample size and diversity of the T2D GWAS, and the new and larger eQTL datasets we employed (**Fig. 2a**). Indeed, the number of colocalizations per eQTL dataset was strongly correlated with the eQTL dataset sample size (Pearson r=0.79, *P*=6.8×10^-3^), highlighting the importance of statistical power in colocalization analyses (**Extended Data** Fig. 1).

**Figure 2:**
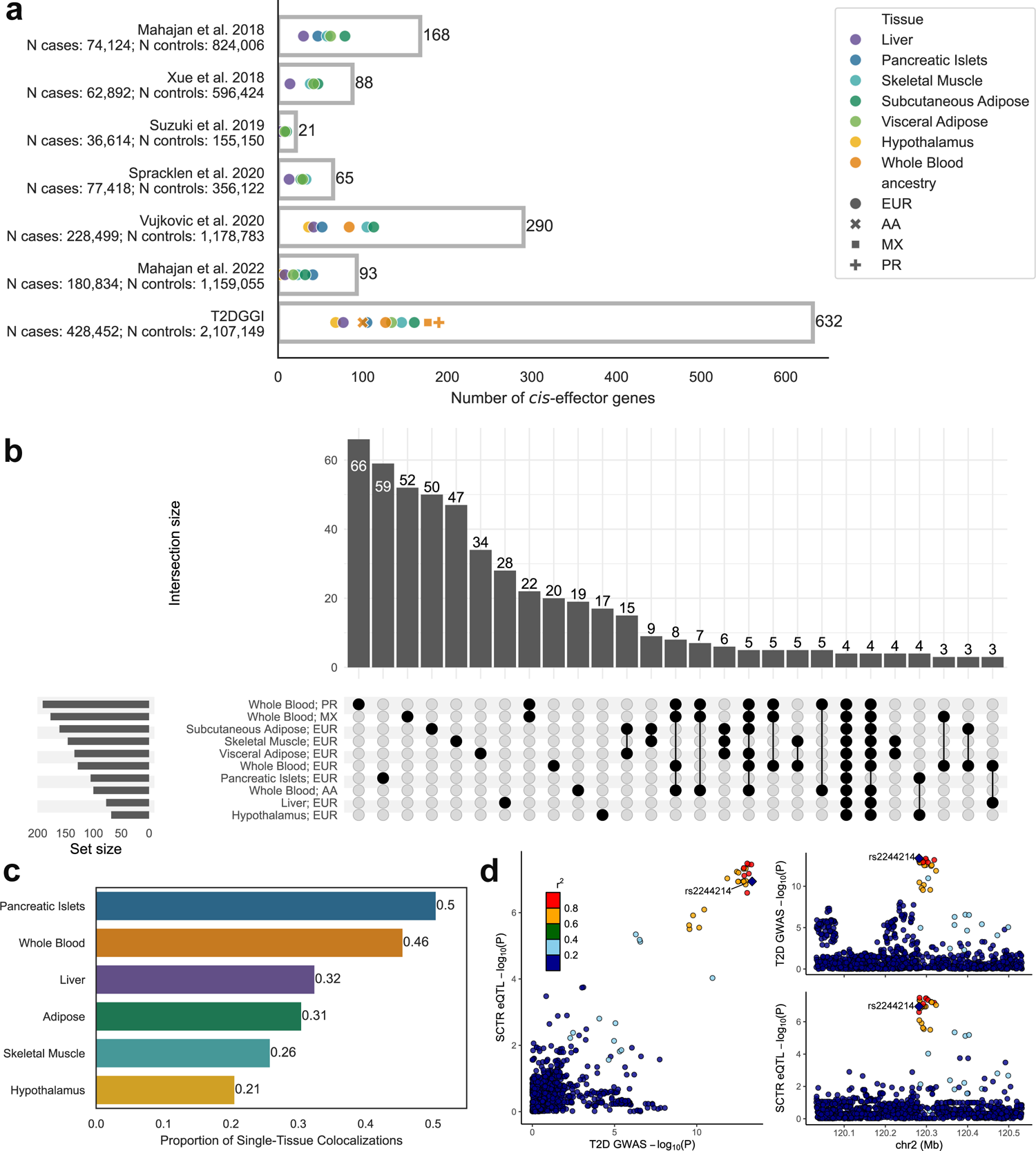
Identification of putative effector genes for T2D. **a)** Plotted are the number of effector genes for T2D previously identified from colocalization analyses between various T2D GWAS and eQTL datasets. Colors indicate the tissue type of the eQTL dataset and shape indicates major self-reported population group of the eQTL dataset. Gray bars represent the total number of unique transcripts across all colocalization analyses per GWAS. **b)** Upset plot of the variant to gene mappings identified in each eQTL dataset analyzed. **c)** Proportion of colocalizations with evidence in one tissue (PP.H4 >0.8) and no positive evidence observed in other tissues (PP.H4 <0.3). **d)** Example of a colocalization observed only in Pancreatic Islets, for the gene *SCTR*. Colors indicate LD in EUR populations from 1000G relative to rs2244214.

We observed many colocalizations in a single tissue only (**Fig. 2b**) with pancreatic islets having the highest proportion of single-tissue observed colocalizations (0.50; **Fig. 2c**). One of the colocalizations only observed in pancreatic islets was with *SCTR,* a G-protein-coupled receptor in the same family as known T2D drug target *GLP1R* (**Fig. 2d**). *SCTR* is highly and specifically expressed in pancreatic islets^19^ and is downregulated in pancreatic islets of T2D patients^25^, supporting the relevance of pancreatic islet expression of *SCTR* in T2D pathophysiology. *SCTR* also harbors a missense variant associated with lower T2D risk^12,13^ that is enriched in EAS-like populations.

To validate the candidate effector genes colocalizing with T2D in islets, we performed gene set enrichment analyses between differentially expressed T2D genes in islets and these effector genes stratified by the direction of effect for the T2D risk allele (**Supplementary Table 8**; **Methods**). We found that genes from colocalizations where decreased expression is associated with increased T2D risk are significantly enriched in genes downregulated in islets of T2D cases (Normalized Enrichment Score [NES]=-1.7; *P*=0.01) (**Extended Data** Fig. 2). Enrichment only in the colocalizations where the risk allele reduces expression may indicate a large abundance of associations between diminished gene function and T2D risk.

### pQTL colocalization identifies additional cis and trans effector proteins

To expand our identification of effector genes beyond eQTLs, we tested for evidence of colocalization between T2D and four pQTL datasets (plasma UKB, plasma deCODE, plasma HELIC, and brain ROS/MAP). We identified 42 and 3,423 colocalizations with a *cis-*pQTL and *trans*-pQTL respectively, encompassing 1,728 unique genes and 365 (28.3%) T2D index variants.

We identified most of the colocalizations (2,451, 70.7%) with pQTLs from the deCODE dataset, which had the second largest sample size but included more proteins, owing to the use of the Somascan panels (N=35,559; N_proteins_=4,907), compared with the OLINK panels employed by the UKB data (N=54,219, N_proteins_=1,472; **Fig. 3a**). We also observed 85 colocalizations with the HELIC data (N=2,933) and 7 with the ROS/MAP data (N=330). Despite differences in sample size and protein sets, we observed 37 identical colocalizations (all in *trans*) with deCODE and UKB pQTL datasets, which is 7.5-fold higher than expected by chance considering the overlap of proteins detected by both the Somascan and OLINK panels (Fisher’s exact test *P*=7.4×10^-20^; **Methods**; **Supplementary Table 9**). Among those 37, we found strong consistency of protein effect size estimates between deCODE and UKB (Pearson r=0.93; *P*=9.1×10^-17^; **Fig. 3b**). One example colocalization detected with both UKB and deCODE pQTL datasets was for CBLN4 with T2D index variant rs1415287 in trans, within the *LYPLAL1* locus defined in Suzuki et al.^15^, where the risk allele associated with increased levels of CBLN4 (**Fig. 3c**). rs1415287 additionally colocalized with IGFBP-1 in *trans* (**Fig. 3d**), with decreased IGFBP-1 levels associated with the T2D risk allele. A previous study found decreased CBLN4 and increased IGFBP-1 plasma levels after administering SGLT2 inhibitors, a class of T2D drugs which reduces glucose reabsorption within the proximal renal tubule, to participants of varying glucose tolerance^26^, mirroring our findings of increased CBLN4 and decreased IGFBP-1 levels with the T2D risk allele.

**Figure 3:**
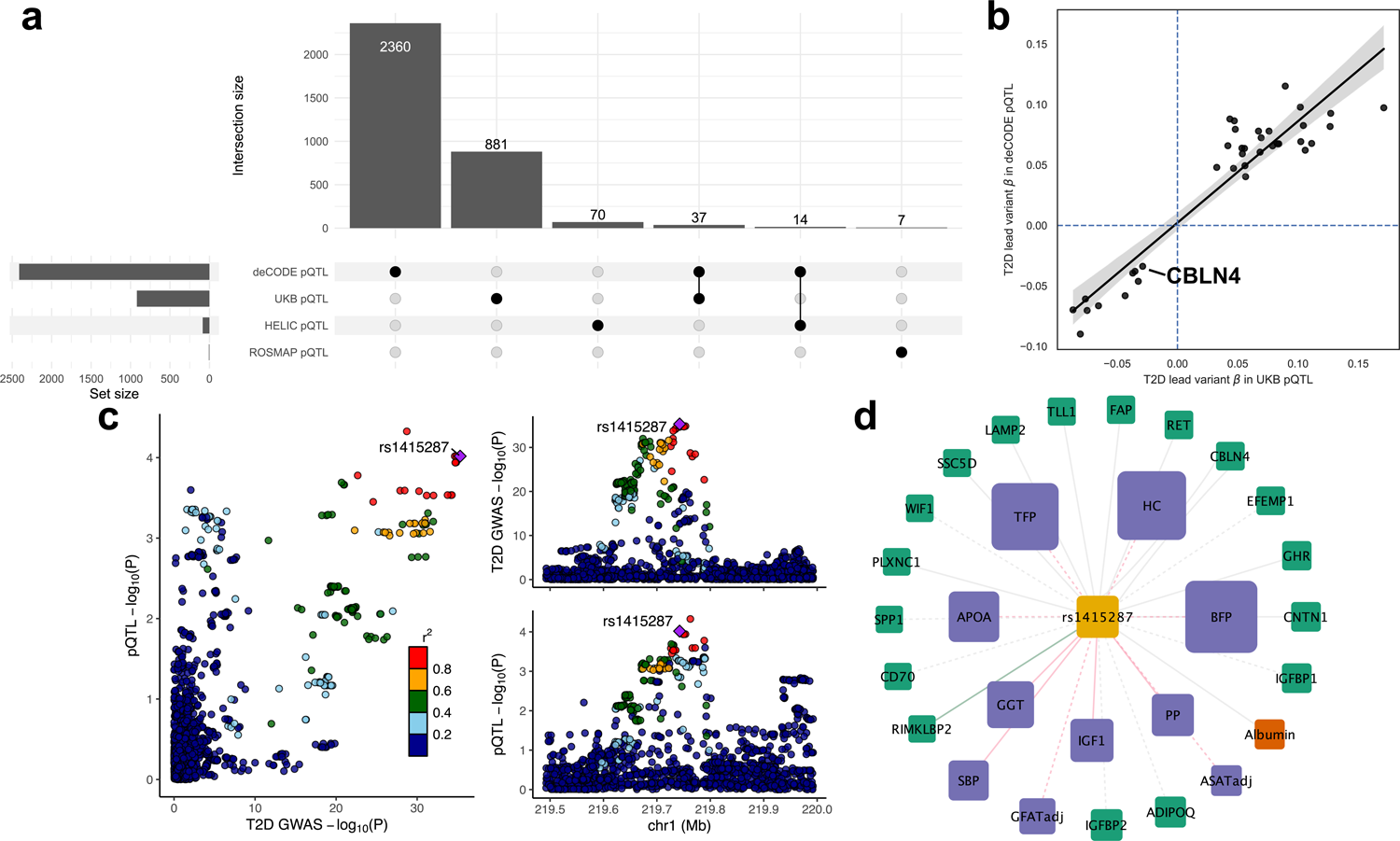
pQTL colocalizations identified in multiple datasets. **a)** Upset plot of variant to gene mappings identified via colocalization analyses with four different pQTL datasets. **b)** Correlation of pQTL effect sizes for colocalizations identified in both UKB and deCODE pQTL datasets (Pearson R=0.93; P=9.1×10^-17^). **c)** Locus compare plot of *CBLN4* using deCODE pQTL data. **d)** Colocalization subnetwork of rs1415287, containing the colocalizations with CBLN4 as well as a colocalization with IGFBP-1. Green nodes represent genes from an eQTL or pQTL, orange nodes represent metabolites, purple nodes represent traits, and yellow nodes represent T2D index variants. Size of the nodes indicate the number of colocalizations observed in the full network. Gray edges represent colocalizations with a plasma/blood dataset, green edges represent colocalizations with a subcutaneous adipose eQTL dataset, and pink edges represent colocalizations with a trait. Dashed lines indicate colocalizations are in the opposite direction as T2D risk and solid lines indicate colocalizations are in the same direction as T2D risk.

We also identified 784 genes for which there was a pQTL colocalizing with more than one T2D index variant (**Extended Data** Fig. 3a). We ranked these genes based on the association between the pQTL and T2D GWAS effect sizes across all index variants that colocalized with a given gene (**Supplementary Table 10**, **Methods**). Among the highest ranked genes, *IGFBP2* had 14 colocalizations with overall negative associations between T2D-associated index variant risk allele effect size and protein levels, supporting previously reported negative associations between IGFBP-2 levels and T2D incidence^27^ (**Extended Data** Fig. 3b). Seven of these index variants (50%) have previously been classified as being involved in lipodystrophy-related pathways^15^, suggesting IGFBP-2 may be involved lipodystrophy-like T2D. In agreement with that, previous publications have found associations between increased IGFBP-2 and lower BMI, lower waist-hip ratio, and increased triglycerides^28^ and decreased NAFLD risk^29,30^. Together, these results highlight the value of highly powered pQTL datasets in identifying biologically relevant effector genes.

### Inclusion of understudied population datasets boosts effector gene identification

In the latest multi-ancestry T2D meta-analysis, 289 index variants (22.4%) have a MAF half as large in EUR-like populations compared to other populations^15^, in total mapping to 615 effector genes from a previously-described T2D-relevant tissue eQTL colocalization or pQTL colocalization. However, identifying effector genes for variants enriched in underrepresented populations remains a challenge due to a lack of molecular data from the same population^31^. To quantify the extent to which including molecular data from underrepresented populations improves the discovery of effector genes, we tested for colocalization between blood eQTL datasets collected in self-reported African American (AA), Mexican American (MX), and Puerto Rican (PR) cohorts^22^ and the EUR-like GTEx dataset with both the multi-ancestry T2D GWAS and with matched ancestry T2D GWAS containing only AFR-like participants within the USA or AMR-like participants. We identified 633 colocalizations with one of these eQTL datasets (47.8% of all eQTL colocalizations), representing 347 gene-to-variant mappings. We identified 204 (58.8%) of the 347 blood eQTL mappings in the AA, MX, or PR eQTL datasets only, and 37 (10.7%) only in the EUR-like GTEx dataset (**Fig. 4a**; **Methods**; **Supplementary Table 11**). In total, adding the AA, MX, or PR eQTL data identified 158 additional T2D effector genes not identified with any of the tested EUR-like eQTL or pQTL datasets, highlighting the importance of collecting data from underrepresented populations, even when testing a tissue that is not directly relevant for T2D pathophysiology. We found that T2D index variants with a colocalization observed only with an AA, MX, or PR blood eQTL were more common in AMR-like participants and less common in EUR-like participants relative to T2D index variants with a colocalization observed only with the EUR-like GTEx blood eQTL dataset (**Fig. 4b**; *P*=0.02). These results suggest differences in statistical power due to allele frequency heterogeneity and/or varying linkage disequilibrium (LD) with the true causal variant may underly differential detection of colocalizations across ancestry groups.

**Figure 4:**
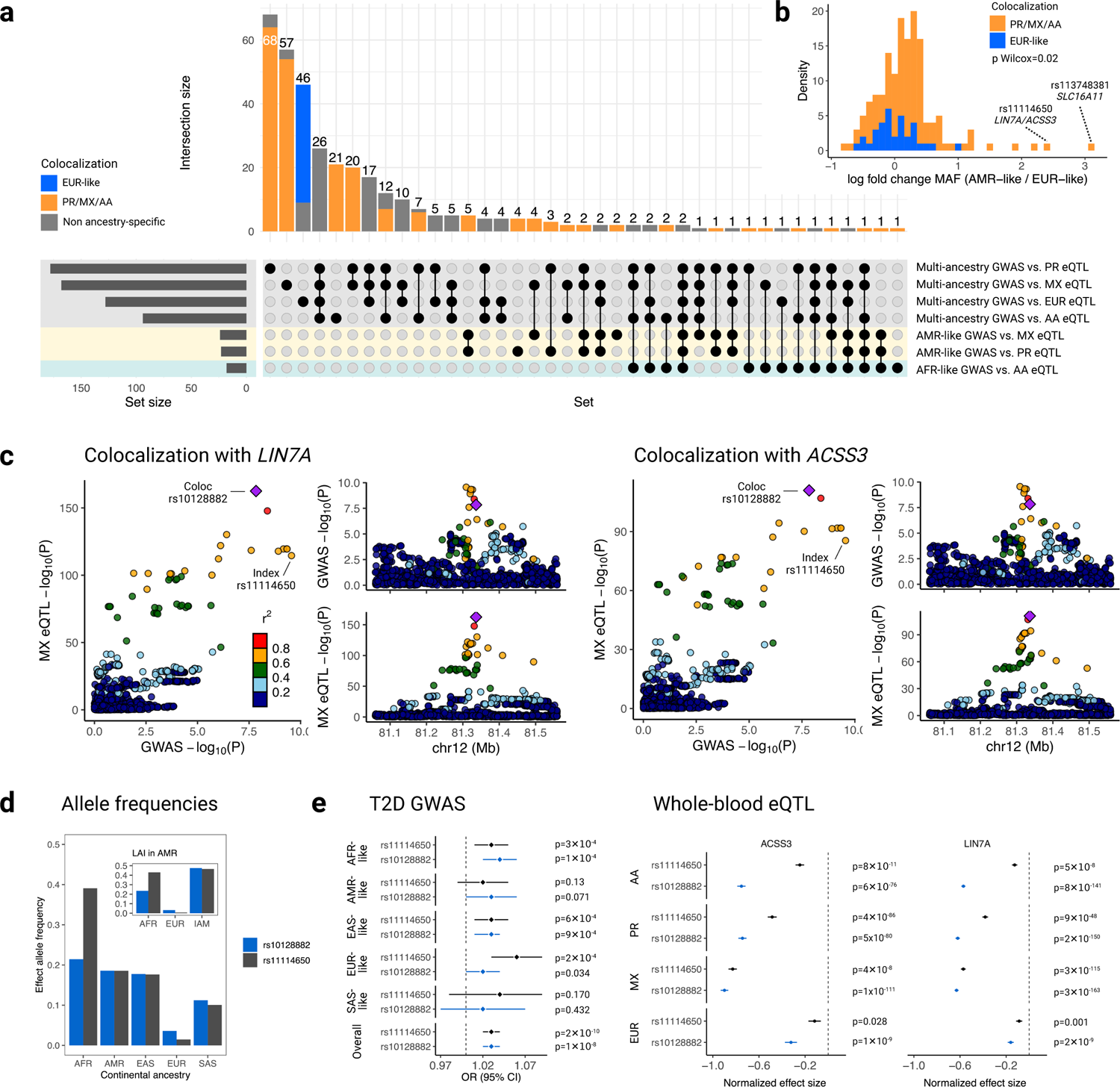
eQTL from understudied populations identify novel colocalizations with T2D. **a)** Upset plot of colocalizations identified with blood eQTL datasets from four different populations. Orange bars represent colocalizations observed only in the Puerto Rican (PR), Mexican American (MX), or African American (AA) datasets (PP.H4 >0.8) and not in the European (EUR) dataset (PP.H4 <0.3). Blue bars represent colocalizations observed only in the EUR dataset (PP.H4 >0.8) and not in PR, MX, or AA datasets (PP.H4 <0.3). Gray bars represent colocalizations observed in any of the PR, MX, or AA datasets and showed a PP.H4 between 0.3 and 0.8 in the European dataset, or were observed in the European dataset but showed a PP.H4 between 0.3 and 0.8 in any of the PR, MX, or AA datasets. **b)** Log-fold change of allele frequencies between AMR-like and EUR-like populations for T2D index variants with a colocalization observed in one population group. **c)** Locus compare plots of *LIN7A* and *ACSS3* with MX data. Colors indicate LD in AMR continental ancestry from 1000G relative to variant rs10128882. **d)** Allele frequencies of lead colocalizing variant rs10128882 (blue) and T2D index variant rs11114650 (black) per continental ancestry and per inferred local ancestry among AMR participants from gnomAD v4.0. **e)** Effect sizes of rs10128882 (blue) and rs11114650 (black) in both the T2D GWAS and blood eQTL datasets, stratified by ancestry.

Two example colocalizations observed only in the MX blood eQTL data were with *LIN7A* and *ACSS3*, both with the same lead variant from colocalization analyses, rs10128882 (**Fig. 4c**). The LD between rs10128882 and the T2D GWAS index variant rs11114650 was different between ancestry groups, with the highest being in EAS-like individuals (r^2^=0.89) and the lowest in AFR-like individuals (r^2^=0.02), possibly reflecting differential LD to the true causal variant across populations. The T2D risk allele of rs11114650 has a frequency of 1.5% in gnomAD-defined European populations compared to 39% in gnomAD-defined African populations (**Fig. 4d**) and is associated with decreased blood expression levels of *LIN7A* and *ACSS3* (**Fig. 4e**). *LIN7A*, mainly expressed in the brain, is involved in localizing, distributing, and maintaining channels and receptors at polarized cell membranes^32^, while *ACSS3* is highly expressed in brown adipose tissue and has been previously associated with insulin-resistant obesity-like phenotypes in mice^33^.

### Identification of effector metabolites

To understand the metabolic processes of T2D-associated genetic variation, we tested for colocalization between T2D GWAS and plasma metabQTL data from UKB and the METSIM study. We identified 5,221 metabQTL colocalizations, corresponding to 283 T2D index variants, with a majority of these colocalizations (4,480, 85.8%) with the UKB metabQTL data. This is expected given the larger sample size of the UKB data (N=114,999) compared to the METSIM data (N=6,136). Of the 18 metabolites measured in the METSIM Metabolon panel that could be mapped to metabolites in the UKB Nightingale panel, 7 colocalized with the T2D GWAS in both METSIM and UKB, an overlap greater than expected by chance (Fisher’s exact test OR=26.2, *P*=1.8×10^-7^; **Methods**; **Supplementary Table 12**), suggesting good concordance between the two platforms for shared metabolites.

To prioritize metabolites with the largest implication with T2D, we ranked metabolites by the number of T2D index variants with which they colocalize, and by the association of genetic effect sizes between T2D and metabolites across all the index variants that colocalized with each metabolite (**Fig. 5a**; **Supplementary Table 13**; **Methods**). We found consistent associations between the risk allele effects and expected metabolite levels, such as T2D risk alleles associated with increased glucose levels (**Fig. 5a**, **5b**). Among less well-established metabolites, we found 10 colocalizations where the T2D risk alleles lowered phosphatidylcholine levels (**Fig. 5c**), consistent with higher phosphatidylcholine intake previously associated with lower risk of T2D^34,35^. A subnetwork of all phosphatidylcholine-linked colocalizations was enriched for metabolites known to be related to phosphatidylcholine metabolism, such as sphingomyelin (*P*_adj._=0.02), cholines (*P*_adj._=2.4×10^-17^), and polyunsaturated fatty acids (PUFAs) (*P_adj._*=0.02; **Fig. 5d**; **Supplementary Table 14**; **Methods**). Within this subnetwork, SMPD1 (sphingomyelin phosphodiesterase 1) protein levels also colocalized with 4 of the same 10 T2D index variants, including two detected in two plasma pQTL studies (UKB pQTL and deCODE pQTL for rs9987289 and UKB pQTL and HELIC pQTL for rs8107974), providing robustness to these results. SMPD1 acts in the same biochemical pathways as phosphatidylcholine synthesis and, along with PUFAs, has previously been associated with diabetic complications including diabetic retinopathy^36^. Combined, these results highlight the role of phosphatidylcholine metabolic pathways in T2D.

**Figure 5:**
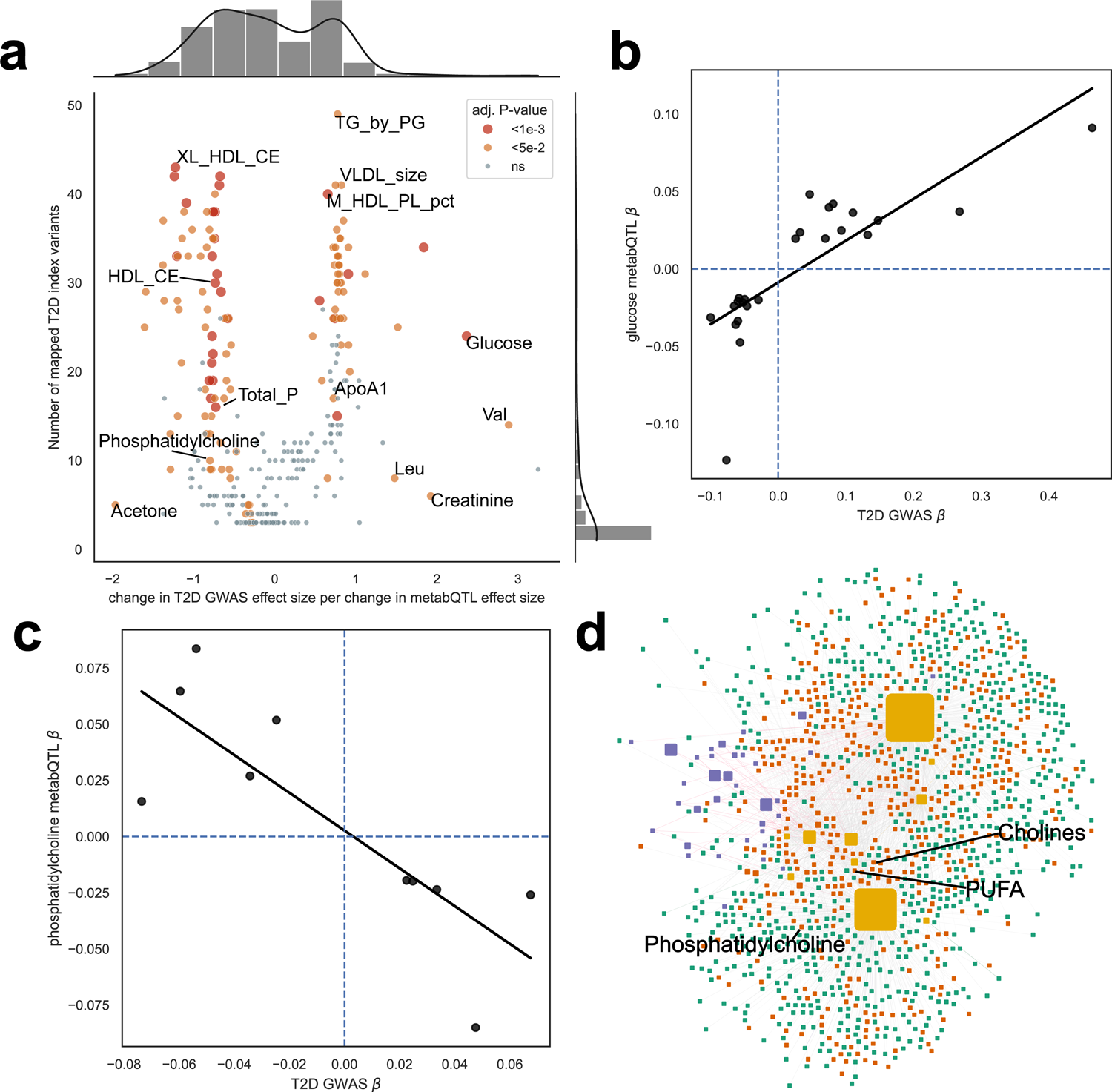
Phosphatidylcholine has consistent negative effect directions with T2D risk. **a)** Joint scatterplot of metabolites comparing the number of T2D-associated index variants they are mapped to compared to the change in T2D effect size per change in metabQTL effect size across the variants. Points are colored and sized by Benjamoni-Hochberg adjusted p-value. **b)** Increased glucose has consistent correlation with increased T2D risk at colocalizing index variants. **c)** Decreased phosphatidylcholine has consistent correlation with decreased T2D risk at index variants. **d)** Subnetwork of all colocalizations mapped to a Phosphatidylcholine-mapped T2D-associated index variant. Green nodes represent genes from an eQTL or pQTL, orange nodes represent metabolites, purple nodes represent traits, and yellow nodes represent T2D index variants. Size of the nodes indicate the number of colocalizations observed in the full network.

### Colocalization with T2D-related traits recapitulates previously described T2D genetic clusters

We next performed colocalization analyses between T2D and 46 T2D-related traits to shed light into distinct T2D pathophysiologic mechanisms. We identified 2,170 colocalizations, including 521 (40.4%) of the T2D index variants and 43 traits.

Previously, Suzuki et al. grouped T2D index variants based on their associations with a smaller subset of 37 of these traits to define 8 distinct genetic T2D mechanistic clusters (Beta cell with positive proinsulin, Beta cell with negative proinsulin, Residual glycaemic, Body fat, Metabolic syndrome, Obesity, Lipodystrophy, Liver/lipid metabolism). Their analyses used a k-means clustering approach on the Z-scores of the T2D index variants from each of the 37 T2D-related trait GWAS^15^. We expanded on this work by assessing the proportion of T2D index variants per cluster which could be mapped to a colocalization between T2D and at least one of the 37 traits previously used for clustering. We found that the Obesity cluster had the highest number of variants confirmed by colocalization, with 163 (70%) of the 223 index variants colocalizing with at least one of the 37 traits, reflecting the large number of obesity-related traits considered during colocalization testing. The colocalizing trait patterns also largely recapitulated the single-variant association-derived clusters^15^ (**Fig. 6a**, left panel).

**Figure 6:**
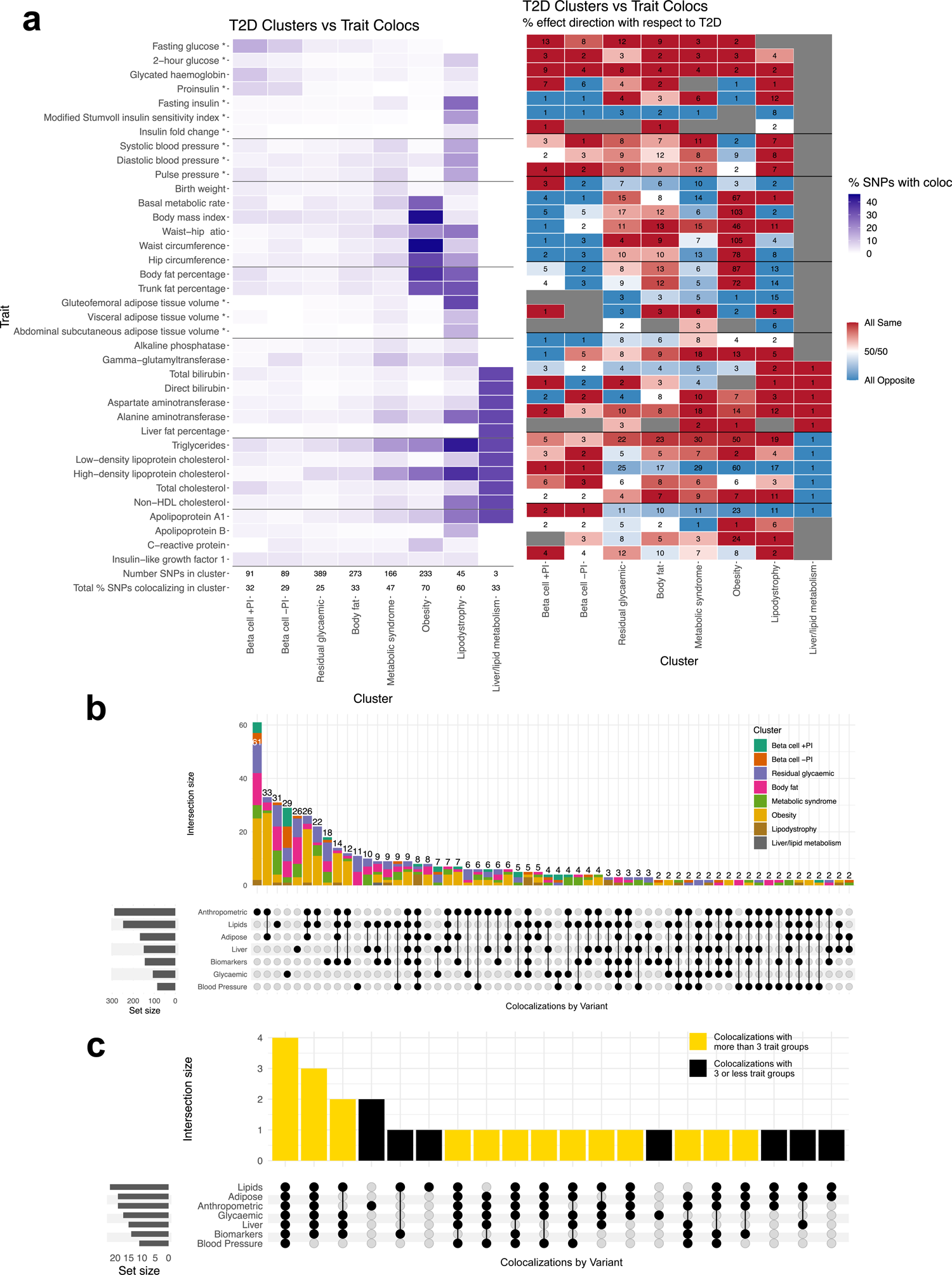
Type 2 Diabetes Clusters Match Trait Colocalizations. **a)** left panel: Color indicates the percentage of total SNPs with a colocalization in each cluster; each row was a trait used for clustering. Final rows list total number of SNPs in each cluster and the total % of SNPs in the cluster with at least one colocalization. **right panel:** Color indicates the percentage of colocalizing SNPs who have the same (red) or opposite (blue) effect direction as the linked T2D SNP, with white indicating that SNPs are split 50/50 for direction, and gray indicating there were no colocalizations between that trait and any SNPs in that cluster. Text in each square indicates the total number of SNPs in that cluster that colocalize with that trait. **b)** Upset plot of all colocalizing SNPs with 2 or more trait group colocalizations showing trait group overlaps, with bars colored by cluster membership. **c)** Upset plot showing trait group overlaps for colocalizing SNPs in the lipodystrophy cluster, with gold bars highlighting overlaps that include 4 or more trait groups (n=19 out of 27 colocalizing SNPs).

We also tested the enrichment of eQTL colocalization from different tissues and metabQTL colocalizations in each of the clusters. We observed enrichment of subcutaneous adipose tissue colocalizations with the Metabolic syndrome cluster (*P*=2.2×10^-5^) and of the pancreatic islet colocalizations with the Beta Cell +PI cluster (*P*=1.6×10^-5^; **Extended Data** Fig. 4; **Supplementary Table 15**). We also observed enrichment of fatty acid metabolites with the Lipodystrophy clusters (*P*=2.0×10^-9^), and of glycolysis related metabolites with the Beta Cell +PI cluster (*P*=4.7×10^-9^; **Extended Data** Fig. 4; **Supplementary Table 16**). When aligning effect allele directions and effect sizes, we again recapitulated cluster patterns from Suzuki et al. based on single variant lookups. The results mirror the effects between Obesity (largely same direction) and Metabolic Syndrome/Lipodystrophy (largely opposite direction) cluster SNPs across anthropometric and adipose traits^15^ (**Fig. 6a**, right panel). As we restricted the analyses to loci that have evidence of colocalization, these results are more specific than those previously presented.

Within each cluster, variants showed different patterns of colocalizations with different traits (**Fig. 6b**). For example, the two Beta Cell clusters have the bulk of their colocalizing variants linked to at least one glycaemic trait, while the variants in the Obesity cluster are linked to anthropometric, adipose, and lipids traits (**Extended Data** Fig. 5). In contrast, the Lipodystrophy cluster variants are more pleiotropic, with 19 of 27 variants linked to 4 or more trait groups, suggestive of heterogeneity within lipodystrophy-related variants (**Fig. 6c**).

### Effector genes are enriched in drug targets for diabetes

The prioritization of drugs targeting candidate genes can accelerate clinical translation^5,37,38^. We pulled known drugs which target one of the 2,311 genes identified from a *cis*-eQTL or a *cis*- or *trans*-pQTL colocalization (**Methods**). Among 1,964 gene-to-drug mappings, consisting of 665 drugs listed as approved, not withdrawn and with information on disease indications, and 159 of our colocalizing genes (**Supplementary Table 17**).

We tested if drugs targeting any of these genes were enriched with an approved indication for diabetes. We observed a 3.5-fold enrichment for approved diabetes indication within drugs that target our list of effector genes (*P*=4.4×10^-10^; **Fig. 7a**; **Methods**) and a 2.0-fold enrichment for any diabetes indication within drugs targeting the effector genes (*P*=1.9×10^-6^; **Fig. 7b**; **Methods**), compared to all other drugs. We then performed the same analysis stratified by the source of colocalization, using lists of 1,681 genes identified only from a pQTLs (*cis* or *trans*), 1,654 only from *trans* pQTLs, or 583 only from eQTLs. Drugs targeting pQTL-only identified genes were enriched for approved T2D drug targets (OR=2.4, *P*=3.4×10^-4^). Drugs targeting eQTL-only identified genes were the most enriched for approved drugs for diabetes (OR=3.9, *P*=3.7×10^-7^). These results suggest that drugs targeting any of our candidate effector genes are potential candidates for repurposing to treat T2D. Permutation analyses similarly found significant enrichment for diabetes drugs among our genes relative to random sets of genes (**Extended Data** Fig. 6; **Methods**).

**Figure 7:**
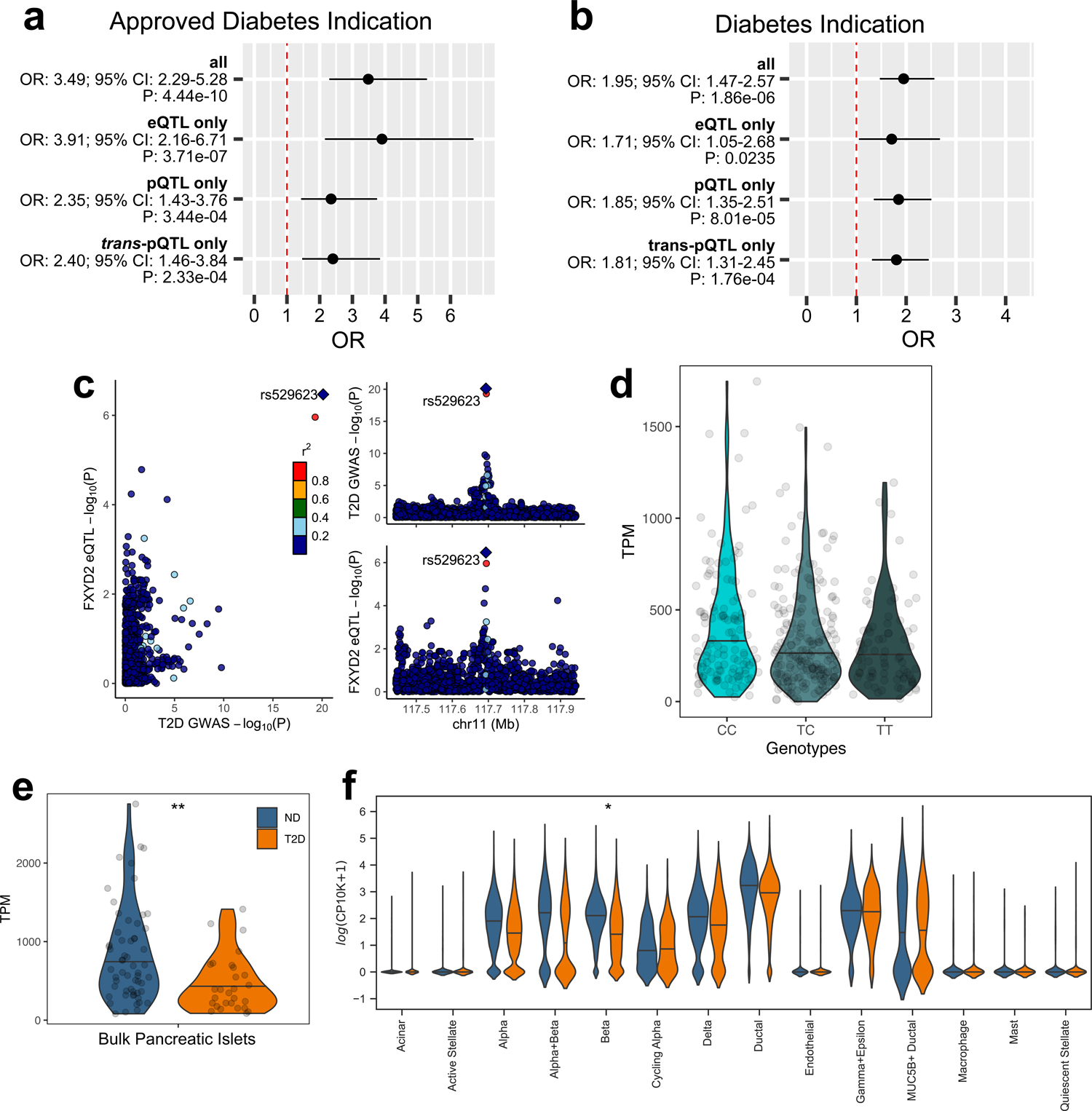
Identification of novel drug targets for T2D. **a)** Forest plot of chi2 enrichment results between drugs targeting colocalizing genes identified in at least one dataset (all), only from an eQTL, only from a pQTL, and only from a *trans*-pQTL with drugs having an approved indication of diabetes in Open Targets. **b)** Forest plot of enrichment results using drugs having an indication of diabetes. **c)** Locus compare plot of *FXYD2*, identified from a colocalization observed only with pancreatic islet eQTL data. **d)** Violin plot of *FXYD2* expression per rs529623 genotype. **e)** Violin plot of *FXYD2* expression among people with T2D and non-diabetes (ND) stratified by weight status in bulk pancreatic islet data. **f)** Violin plot of *FXYD2* expression from human donor single-cell pancreatic islet data stratified by cell-type. Adjusted P-values <0.001 are indicated with “**”, and adjusted P-values <0.05 with “*”. Due to the low number of cells per sample, the differential expression test was not performed for some cell types (**Methods**).

Among the genes observed only from an eQTL colocalization, we identified *FXYD2* (**Fig. 7c**) which has been proposed as a pancreatic beta-cell specific biomarker^39^. FXYD2 modulates Na,K-ATPase activity and cell proliferation^40^ and is expressed at the early stages of the human endocrine pancreas development (15 weeks), preceding insulin detection^39^, suggesting a putative role in islet development. We also detected *FXYD2* mRNA expression at the end of the differentiation process of induced pluripotent stem cells into islet-like cells in both bulk and single-cell datasets^28,41–43^ (**Extended Data** Fig. 7).

The risk allele of the T2D index variant rs529623 was associated with lower expression of *FXYD2* in pancreatic islets^19^ (**Fig. 7d**; *P*=3.4×10^-7^). We further explored expression differences of *FXYD2* in existing bulk RNA-seq^44^ and single-cell RNA-seq^45^ (scRNA-seq) from pancreatic islets of human donors with and without T2D (**Methods**). After adjusting for multiple hypothesis testing, *FXYD2* was significantly downregulated in pancreatic islets of T2D patients (**Fig. 7e**) specifically within Beta cells^45^ (**Fig. 7f**). There are currently five approved drugs that target FXYD2 but are not approved for diabetes: delanoside, digoxin, digitoxin, acetyldigitoxin, and lanatoside. These drugs potentially could be modified to treat T2D, particularly if further preclinical investigations show indications that they can target and modulate beta cell function.

We also found multiple other drugs targeting proteins for one of our effector genes which had clinical evidence to support a role in impacting glycemia and may represent possible targets for drug repurposing (**Supplementary Table 18**). For example, amisulpride, an antagonist for the Serotonin (5-HT) receptor encoded by *HTR6*, increases pancreatic insulin secretion in healthy controls^46^. We identified *HTR6* from a blood *trans-*pQTL colocalization where the T2D risk allele is associated with increased protein levels, consistent with amisulpride, which antagonizes HTR6 and increases insulin secretion.

## Discussion

T2D is a model for complex disease genetics owing to its high prevalence and polygenicity, with the largest T2D multi-ancestry meta-analysis to date based on >2.5 million participants, identifying 1,289 index variants^15^. However, understanding the function of associated variants from a GWAS is extremely challenging, and only few examples have provided additional insights into the molecular mechanisms underpinning signals^47–51^. These comprehensive studies take years to complete, and biological insights from previous studies were mostly described in EUR-like populations^14,18,19,24^, limiting findings for other populations in which T2D risk alleles are now being increasingly reported^11,15^.

Here, we aimed to create a catalog of effector genes, metabolites, and traits for the 1,289 T2D-associated index variants. We identified >12,000 colocalizations with T2D across all omics layers and T2D associated traits tested. We found *cis*-effector genes for 361 of the index variants (28%) and identified a >2-fold increase in the number of *cis-*effector genes compared with the previous largest T2D GWAS^14^. We observed many of the eQTL colocalizations in only one of the tissues tested, emphasizing the importance of including multiple tissue types in variant-to-function efforts for T2D. We also demonstrated the value of multi-ancestry eQTL data through the identification of 158 additional effector genes (24% of all *cis-*effector genes) using blood eQTL data from AA, PR, and MX populations^22^ that we did not observe in eQTL or pQTL data from EUR-like populations alone. We demonstrate that even in blood, which is not directly related to T2D progression, including eQTL data from multiple ancestry groups improves effector gene discovery. Despite substantial progress in the inclusion of diverse ancestries in GWAS meta-analysis, the availability of molecular data from diverse ancestries remains limited^31^. We hope that the present findings will help motivate the generation of molecular QTL resources from participants of diverse ancestries to unveil additional effector genes and potential drug targets.

We compiled the pairwise colocalizations between T2D and each omics layer and additional traits to create a network of T2D index variants mapped to genes, metabolites, and traits. This provides the most comprehensive resource to date of the genetic relationships between genes, traits, metabolites, and genetic variants associated with T2D to the research community. Such a resource allows searches for genes, metabolites, or traits of interest and find the T2D index variants underlying their shared causal signals with T2D to quickly generate hypotheses and gain insights into T2D pathophysiology. We additionally include effect and variant information from the molecular QTL, trait GWAS, and T2D GWAS summary statistics to further improve the searchability of the network.

Identifying drug targets for complex diseases has traditionally focused on protein coding variants, which involves designating the target gene and mimicking a loss-of-function or gain-of-function effect^52,53^. However, effector genes regulated by non-coding variants may also be viable drug targets once the mechanism of action is understood. In this study, we identified candidate effector genes, their tissue of action and direction of effect, for potential drug target identification. We show that our list of effector genes is enriched for targets of approved drugs for treatment of T2D, suggesting that drugs not currently used to treat diabetes but targeting one of the effector genes are strong candidates for modification for the treatment of T2D. We acknowledge that additional studies to test causality, druggability, and functional validation with tissue and cell types of interest are needed.

Our study has limitations. First, we utilized the 1,289 index variants from Suzuki et al. to define our tested colocalization regions, but we recognize that these variants were not assessed for statistical independence applying formal fine-mapping approaches. Second, we leveraged the multi-ancestry meta-analysis for T2D to maximize sample size. However, this approach may underestimate the true number of colocalizations, as we could not perform fine-mapping to differentiate multiple independent signals within the same locus due to current limitations in fine mapping and conditional analyses utilizing only multi-population summary statistics data. Third, confirmation of the tissue of action and the causality and directionality of the effect, especially for distal-effector genes, remains to be determined. This necessitates careful investigation through appropriate causal inference experiments, cellular models, and *in vivo* studies. To facilitate these efforts, we provide here the most extensive list of effector genes, tissues, and potential physiological pathways linked to T2D associated variation for initial prioritization strategies.

In conclusion, we present a comprehensive functional follow-up of the largest T2D GWAS meta-analysis and provide a framework model for functional annotation of other complex multi-ancestry GWAS. We provide an interactive network to explore the relationships between T2D-associated variants and genes, metabolites, and traits. We anticipate that this resource will serve as a valuable tool for generating testable hypotheses that may lead to the discovery of novel T2D drug targets and therapeutic strategies.

## Methods

### GWAS and Molecular Trait Associated Datasets

We downloaded GWAS for T2D relevant traits downloaded from a variety of sources described in **Supplementary Table 1**^54–70^. We used T2D multi-group meta-analysis generated using METAL^71^ from Suzuki et al.^15^ for all colocalization analyses. Additionally, single population group meta-analyses using METAL from Suzuki et al. for AFR-like participants living within the USA and for AMR-like participants were used for colocalization. We removed variants with a MAF <0.5% across all the 1000G-continental similarity groups from the analyses.

We downloaded pancreatic Islet eQTL data from TIGER^19^. Since effect size and variance information is not available in the TIGER eQTL data, we used p-value information from the summary statistics and allele frequencies from UKB to calculate approximate bayes factors for colocalization analyses. We downloaded eQTL data for subcutaneous adipose, visceral adipose, skeletal muscle, hypothalamus, and liver tissue and blood from GTEx v8. We also tested eQTL data for whole blood from self-reported African American (AA), Mexican American (MX), and Puerto Rican (PR) populations for colocalization^22^. Additionally, we tested T2D GWAS including only AFR-like participants from the USA for colocalization with the AA blood eQTL data and tested T2D GWAS including only AMR-like participants for colocalization with the MX and PR blood eQTL datasets. We restricted the colocalization analyses to just these eQTL datasets as pancreatic islets, subcutaneous adipose, visceral adipose, skeletal muscle, hypothalamus, and liver are known to be relevant to T2D. Despite blood representing multiple tissues and not being a target of T2D, the availability of blood eQTL from diverse populations motivated their inclusion.

We performed integration with pQTL data using summary statistics from deCODE, UKB, HELIC, and ROS/MAP. The deCODE pQTL study contains 4,907 aptamers measured with the SomaScan version 4 assay (SomaLogic) in the plasma samples of 35,559 Icelanders^72^. The UKB Pharma Proteomics Project characterized 2,941 plasma protein analytes of 54,219 UKB participants using the antibody-based Olink Explore 3072 proximity extension assay and tested for pQTL with 2,923 proteins^73^. pQTL for only 1,472 of these proteins were available at the time of this study. Plasma pQTL from the Hellenic Isolated Cohorts (HELIC)^74^ included 2,933 samples from two isolated Greek populations (Pomak and the Mylopotamos villages) and 543 proteins from six different OLINK panels: Cardiovascular II, Cardiovascular III, Metabolism, Neurology, Neuro-exploratory and Cardiometabolic^75–77^. Genetic and proteomic data from post-mortem samples of the dorsolateral prefrontal cortex of older adults was provided by the Religious Orders Study (ROS) and Rush Memory and Aging Project (MAP)^78^. The pQTL analysis was conducted for every HapMap3 common variant (MAF >0.05) within a 100 Kb window around protein-coding genes.

For the colocalization analyses with metabolites, we used summary statistics from the METSIM and UKB cohorts. The METSIM metabolomics study measured 1,391 plasma metabolites in 6,136 Finnish men using the Metabolon Discovery HD4 mass spectrometry platform^79^. In the UKB cohort, a total of 249 metabolites (168 absolute levels and 81 derived ratios and percentages) were measured from the Nightingale panel and tested for genome-wide associations with 114,999 individuals of European ancestry^80^.

### Colocalization Analyses

We performed pairwise colocalizations between the multi-group T2D GWAS meta-analysis and eQTL, pQTL, metabQTL, and relevant trait GWAS datasets using the *coloc.abf* function from the *coloc*^81^ package in R. *coloc.abf* requires regions to be specified to test for colocalization. To define these regions, index variants were sorted by p-value and a 500 Kb window was constructed around each variant. If a less significant index variant was within the window of a more significant variant, the less significant variant’s window was removed from the list of regions. To prevent additional overlap of regions, any regions overlapping by more than 200 Kb were merged and regions overlapping by less than 200 Kb were split by half of their overlap size. The final region boundaries used in the analyses can be found in **Supplementary Table 2**. A colocalization analysis was performed between a T2D region and a QTL or cardiometabolic trait if and only if there was evidence of an association (defined as P <1×10^-4^) of the T2D index variant or a proxy for the T2D index variant for that trait. Proxy variants were defined as a variant in strong linkage disequilibrium (LD, r^2^ >0.8) in all continental population groups defined by in 1000G (AFR, AMR, EAS, EUR, and SAS).

*coloc.abf* provides the posterior probability that a region shares a causal variant in both association summary statistics tested and posterior probabilities that each variant present in both summary statistics is the causal variant. A colocalization was deemed of interest if the posterior probability or shared variation (PP.H4) was greater than 0.8. To determine if a T2D GWAS index variant is the most likely shared causal variant for a region in two colocalizing datasets, the variant with the highest posterior probability of being shared was mapped to a index variant if it was in partial-to-strong LD with the index variant (r^2^ >0.5) in 1000G population groups described above.

We labelled an eQTL colocalization as being observed in one tissue or population group dataset if the PP.H4 was greater than 0.8 in the dataset, but less than 0.3 in all other tissue or population group datasets. A maximum of 0.3 was chosen to avoid selecting suggestive colocalizations that may exist but not be sufficiently powered to be detected with our datasets.

For single tissue analyses, we combined subcutaneous adipose and visceral omentum adipose results and combined blood results from each ancestry due to the larger similarities in gene expression profiles between these datasets relative to the other tissues analyzed.

We characterized pQTL colocalizations as being in *cis* if the mapped index variant was within 1 Mb of the pQTL gene TSS; otherwise they were denoted as in *trans*. We tested for enrichment of colocalizations identified between deCODE pQTL which uses the Somascan assay and UKB pQTL which uses the Olink assay, using Fisher’s exact test. We constructed contingency tables from all gene to T2D index variant mappings possible for genes included in Somascan and/or Olink respectively. We also used this approach to test for an enrichment of colocalizations identified between METSIM metabQTL and UKB metabQTL, using all metabolite to T2D index variant mappings possible for metabolites included in METSIM and/or UKB respectively.

We also tested for enrichment between T2D index variants mapped to an eQTL tissue colocalization and T2D index variants within different T2D genetic clusters using Fisher’s exact test. We constructed contingency tables based on whether one of the 1,289 T2D index variants is within a specific T2D genetic cluster and is also the lead colocalizing variant for a colocalization with an eQTL dataset. The Liver/Lipid Metabolism cluster was excluded due to only having 3 total T2D index variants. We then adjusted P-values using the Benjamini-Hochberg procedure. We repeated these enrichments analyses for UKB metabQTL colocalizations, grouped based on metabolite class.

### Colocalization Network

We constructed a network of all the colocalizations with the T2D meta-analyses using cytoscape^82^ and measured network properties using cytoNCA.

### Effect Direction Consistency

We mapped many of the genes and metabolites identified from a pQTL or metabQTL dataset to multiple T2D index variants. We then measured the consistency of effect estimates for T2D and the pQTL or metabQTL for genes or metabolites with at least 3 mapped T2D index variants by testing for an association between pQTL or metabQTL effect sizes with T2D effect sizes using an ordinary least squares regression model without an intercept, adjusting for an FDR of 0.05 using the Benjamini-Hochberg procedure.

### Expression Dataset Preparation

We downloaded scRNA-seq data from HPAP^45^ analyzed it with Scanpy^83^ v1.9.3. The published data from HPAP we used in this study was pre-QC’ed, clustered, and annotated, and a full description of the HPAP data processing steps can be found in their publication^45^. Briefly, cells were filtered to have >500 expressed genes per cell and <15% mitochondrial reads. After, ambient RNA was adjusted using SoupX, followed by batch correction with Harmony and clustering using the Leiden algorithm with a resolution of 0.5. Cell types were defined by pseudo-bulking each cell type and identifying marker genes identified with a Wald test. We subset cells from healthy and T2D donors and created pseudobulks per sample by summing raw counts across cells per cell-type. We only kept pseudobulks with at least 5×10^5^ counts and from adult individuals (age >= 20 years; N=21 for healthy and N=17 for T2D). We performed the DE analysis with edgeR^84^ v3.40.2. We removed lowly expressed genes per cell-type with filterByExpr, which removes genes with less than 10 counts in 10 samples or 15 counts in all samples, using diabetes status as a stratification covariate. After filtering, we tested a total of 9,995 genes in acinar cells, 8,150 genes in active stellate cells, 13,171 genes in alpha cells, 13,095 genes in beta cells, and 9,422 genes in ductal cells. We then fit a generalized linear model with robust dispersion to the data, using as covariates diabetes status, age, sex, sex and age interaction, self-reported race and ethnicity, and sequencing chemistry. We next used the glmLRT function to identify T2D differentially expressed genes incorporating the dispersion estimates and adjusted the p-values with the Benjamini-Hochberg procedure.

We pulled bulk expression dataset for human islets from Marselli et al.^44^, including 28 T2D cases and 58 ND controls. We performed transcript quantification using salmon 1.4.0^85^, and differential expression tests using DESeq2 1.38.3^86^. The DESeq2 results from this study are accessible on GEO under accession number GSE159984.

We downloaded bulk expression data of *FXYD2* from different iPSC differentiation stages from GEO under the accession number GSE190727. We also downloaded single-cell expression data for stem-cell-derived endocrine cells from Krentz et al. 2018^42^, Xin et al. 2018^28^, and Balboa et al. 2022^43^.

### Open Targets Drug Identification

To identify potential drug repurposing options from our candidate gene list, we queried data files from Open Targets^87^ on around 7k drug molecules and around 62k targets, where 4,930 drugs were annotated with 1,538 gene targets. After also retrieving the reported indications of drugs, we refined our search to the approved drugs, which were not withdrawn and did list indications. Open Targets used drug annotations from ChEMBL to define drugs as being approved for an indication if they come from a source of approved drug information (e.g. FDA, WHO ATC, EMA, BNF). To test if diabetes drugs were identified more often than expected by chance, we compared the drugs targeting our query genes and those targeting all documented non-query genes using chi-squared tests. Additionally, we tested if permuted 100k random samples from the same gene list would identify diabetes drug targets at the same rate, sampling 42,539 known genes with HGNC gene symbols and comparing the results from our query gene set and the random samples.

## Data Availability

All data produced in the present work are contained in the manuscript

## Acknowledgement

A.H. is supported by the American Diabetes Association grant #11-23-PDF-35. O.B. has received funding from the European Union’s Horizon 2020 research and innovation programme under Grant Agreement No 101017802 (OPTOMICS). A.P. is supported by the Wiener-Anspach Foundation, and the Fonds National de la Recherche Scientifique (FNRS). J.B.M is supported by NIDDK (U01DK078616 and R01DK078616). J.M.M. is supported by American Diabetes Association Innovative and Clinical Translational Award 1-19-ICTS-068, American Diabetes Association grant #11-22-ICTSPM-16, by NHGRI (U01HG011723), NIDDK (UM1-DK078616) and Medical University of Bialystok (MUB) grant from the Ministry of Science and Higher Education (Poland). This study also supported in part by the National Center for Advancing Translational Sciences, CTSI grant UL1TR001881, and the National Institute of Diabetes and Digestive and Kidney Disease Diabetes Research Center (DRC) grant DK063491 to the Southern California Diabetes Endocrinology Research Center. Infrastructure for the CHARGE Consortium is supported in part by the National Heart, Lung, and Blood Institute (NHLBI) grant R01HL105756. This research was conducted in part using data and resources from the Framingham Heart Study of the National Heart Lung and Blood Institute of the National Institutes of Health and Boston University School of Medicine. The Framingham Heart Study (FHS) acknowledges the support of contracts NO1-HC-25195, HHSN268201500001I and 75N92019D00031 from the National Heart, Lung and Blood Institute and grant supplement R01 HL092577-06S1 for this research. We also acknowledge the dedication of the FHS study participants without whom this research would not be possible.

## Disclosures

J.B.M is an academic associate for Quest Diagnostics Inc. Endocrine R&D. MIMcC is now an employee of Genentech and a holder of Roche stock.

## Contributions

R.M. Performed eQTL colocalization analyses, integrated all the colocalization results, built the omics network, and characterized the network. Lead the writing of the manuscript and design of figures. Lead the interpretation and discussion of the results, deep dives into specific locus and wrote the paper.

K.M.L. Contributed to design the colocalization pipeline. Performed colocalization analyses with GWAS traits. Integrated the colocalization results with the previously described T2D genetic clusters. Contributed to interpretation and discussion of the results, deep dive into specific locus, design of the figures, discussion of the narrative and writing of the paper.

X.Y. Performed colocalization analyses with deCODE pQLT, UKB pQTL, METSIM metabQTL. Contributed to interpretation and discussion of the results, deep dive into specific locus, design of the figures, discussion of the narrative and writing the paper.

O.B. Performed colocalization analyses with UKB metabQLT. Supervised the enrichment analyses of drug targets using open targets data. Contributed to interpretation and discussion of the results, deep dive into specific locus, design of the figures, discussion of the narrative and writing of the paper.

A.H. Performed colocalization analyses with blood eQTLs from diverse ancestries. Identified and evaluated the contribution of eQTL data from diverse ancestries in identifying effector genes as well as the value of using ancestry-specific GWAS meta-analysis in improving the identification of effector genes. Contributed to interpretation and discussion of the results, deep dive into specific locus, design of the figures, discussion of the narrative and writing the paper.

A.L.A Performed colocalization analyses with ROSMAP and HELIC pQLT. Contributed to interpretation and discussion of the results, deep dive into specific locus, design of the figures, discussion of the narrative and writing the paper.

A.P. Performed the analysis of differential expression in pancreatic islets in T2D and controls and islets in lipoglucotoxic conditions. Performed enrichment analyses of colocalization genes with differentially expressed genes. Contributed to interpretation and discussion of the results, deep dive into specific locus, design of the figures, discussion of the narrative and wrote the paper.

S.H. Performed the curation and mapping of effector genes to drugs using open targets data.

Performed the enrichment analyses of effector genes with drug targets. Contributed to interpretation and discussion of the results, design of the figures, discussion of the narrative and writing of the paper.

K.Y, K.Hr, Y.T., M.L., H.L, M.C, D.L.E. provided and analyzed functional genomics data related from single-cell and pancreatic islets with the FXYD2 locus. L.S., K.S., K.Ha., H.J.T, N.W.R., J.B.M, MIMcC, A.M., M.U., C.N.S, M.B, M.V, J.I.R. and A.P.M Provided feedback throughout the project and revised the paper. R.M., A.P.M, B.V., E.Z and J.M.M conceived and planned the study, supervised and coordinated the analyses, and wrote the manuscript.

**Extended Data Figure 1: eQTL colocalizations correlate with sample size.** Scatter plot of the number of colocalizations between the T2D multi-ancestry meta-analysis and different eQTL datasets compared to the sample size of the eQTL datasets.

**Extended Data Figure 2: Enrichment of pancreatic islet colocalizing effector transcripts in differential gene expression data.** GSEA plots of colocalizing effector transcripts with differentially expressed genes from the bulk human pancreatic islet dataset, stratified by whether the T2D index variant risk allele is associated with increases (same direction) or decreases (opposite direction) of gene expression.

**Extended Data Figure 3: IGFBP2 associations with T2D are highly consistent genome-wide. a)** Joint scatterplot of proteins comparing the number of T2D-associated index variants they are mapped to compared to the association of pQTL effect sizes to T2D effect sizes across the variants. Points are colored and sized by Benjamoni-Hochberg adjusted p-value. **b)** Decreased plasma protein levels of insulin growth factor binding protein 2 *IGFBP2* has consistent correlation with increased T2D risk at colocalizing lead variants.

**Extended Data Figure 4: Overlap of T2D clusters with trait, eQTL, and metabQTL colocalizations.** Heatmaps of the percent of index variants per cluster mapped to a colocalization with a metabolite group, eQTL dataset, and additional related trait GWAS. **Extended Data** Figure 5**: Colocalizations between index variants and trait groups per T2D genetic cluster.** Upset plots of colocalizations between T2D index variants from different T2D genetic clusters with related cardiometabolic trait groups.

**Extended Data Figure 6: Permutations of overlaps from T2D colocalizing genes with drugs from Open Targets.** Ratios of approved diabetes drugs of detected drugs compared to 100k random samples (histogram) for a) genes from all T2D colocalizations, b) genes only identified from a T2D coloclization with eQTL, c) genes only identified from a T2D coloclization with pQTL, and d) genes only identified from a T2D coloclization with *trans*-pQTL. P-values indicate difference in the approved proportion of diabetes indications per drug for colocalizing query genes relative to random permutations.

**Extended Data Figure 7: *FXYD2* expression in islet progenitor cells. a)** Heatmap of *FXYD2* expression throughout various stages of iPSC differentiation into beta-like cells, where colors correspond to the log_2_(TPM+1) expression level of an individual sample (1-5). The differentiation stages include definitive endoderm (DE), pancreatic progenitor (PP), stem cell-derived islets at stage 7 (SC-islets) and the islets following grafting into immune deficient mice for 4 months (graft). **b)** Dot plot showing mean gene expression of FXYD2 and INS in the endocrine cell clusters in the integrated dataset of 46,261 stem-cell-derived endocrine cells and adult human islet cells (Krentz et al. 2018, Xin et al. 2018, and Balboa et al. 2022). Dot size is relative to the fraction of cells within a cluster expressing the gene. Endocrine Prog., Endocrine progenitors; SC-EC, Stem-cell-derived enterochromaffin-like cells; SC-Beta, Stem-cell-derived beta cells; SC-Alpha, Stem-cell-derived alpha cells.

## Notes

### Author Declarations

The study used ONLY openly available human summary statistics data described in Supplementary Table 1 and human gene expression data downloaded from GEO accession number GSE159984 and the Islet Expression HPAP project.

