## Supplementary figures and images for "Multi-omics characterization of type 2 diabetes associated genetic variation"

### Extended Data Figure 1

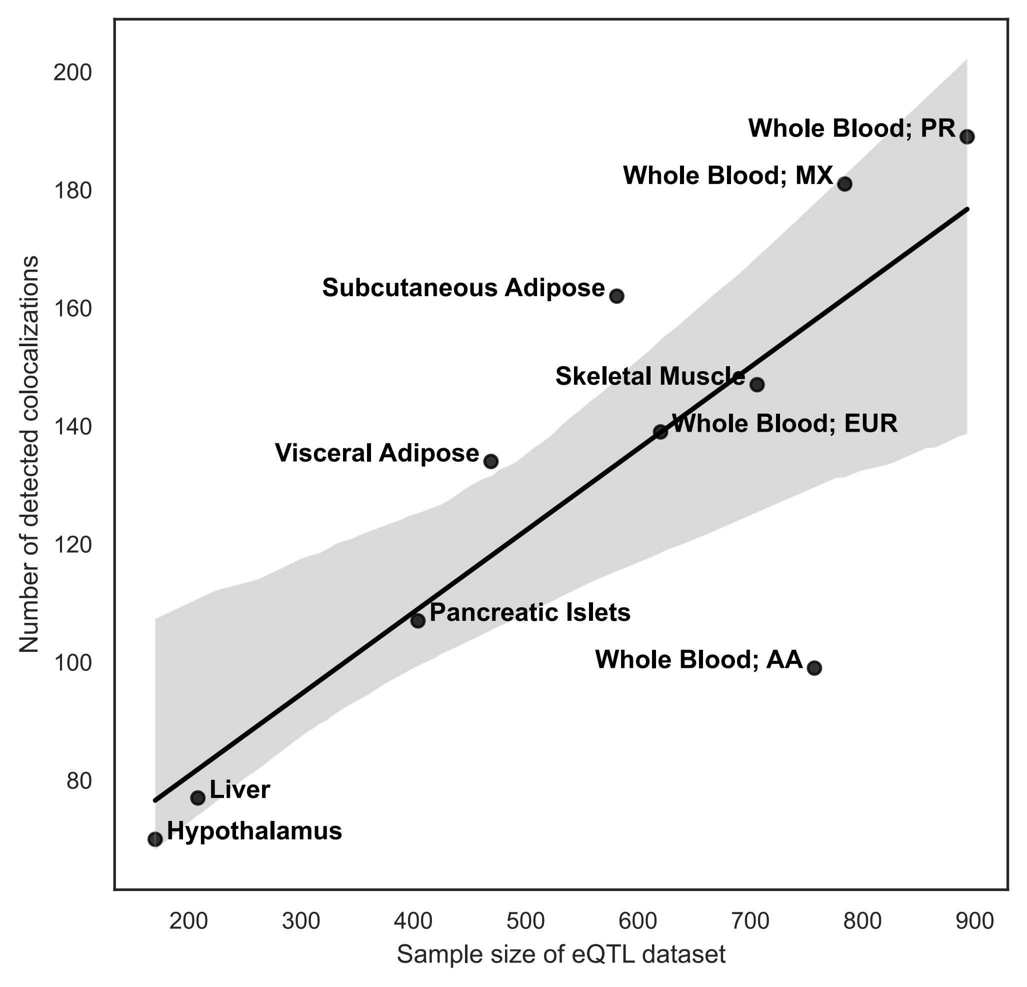

### Extended Data Figure 2

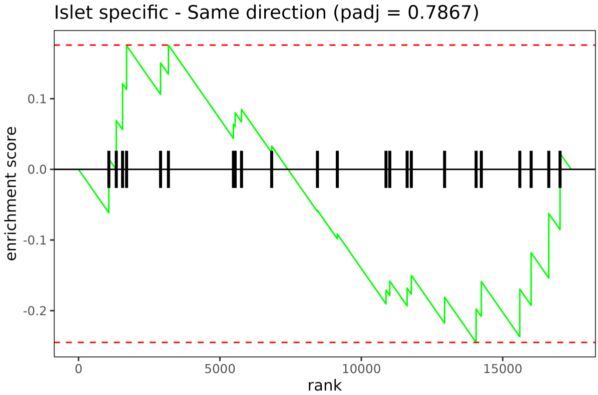

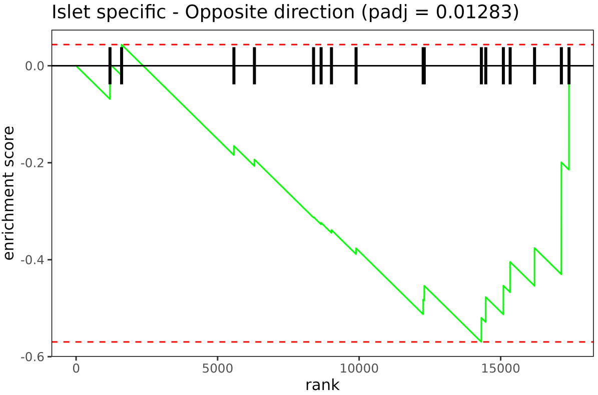

### Extended Data Figure 3

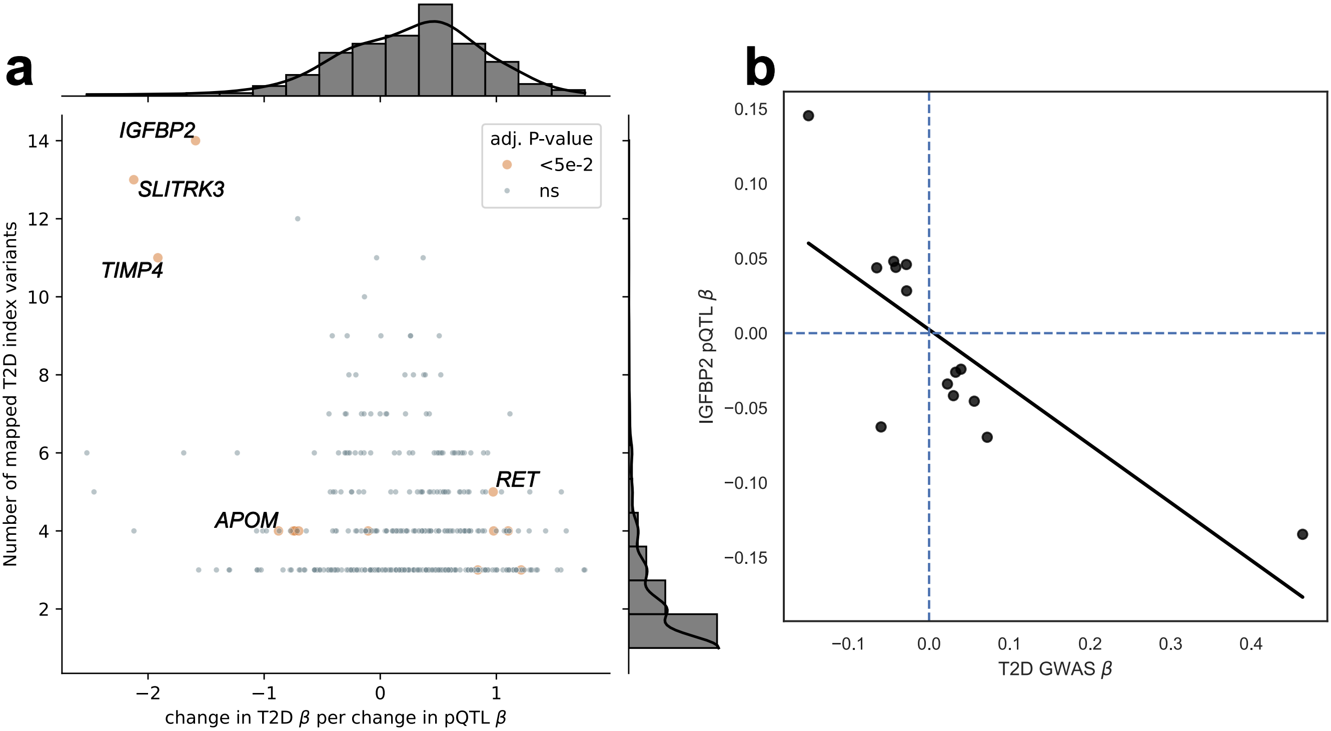

### Extended Data Figure 4

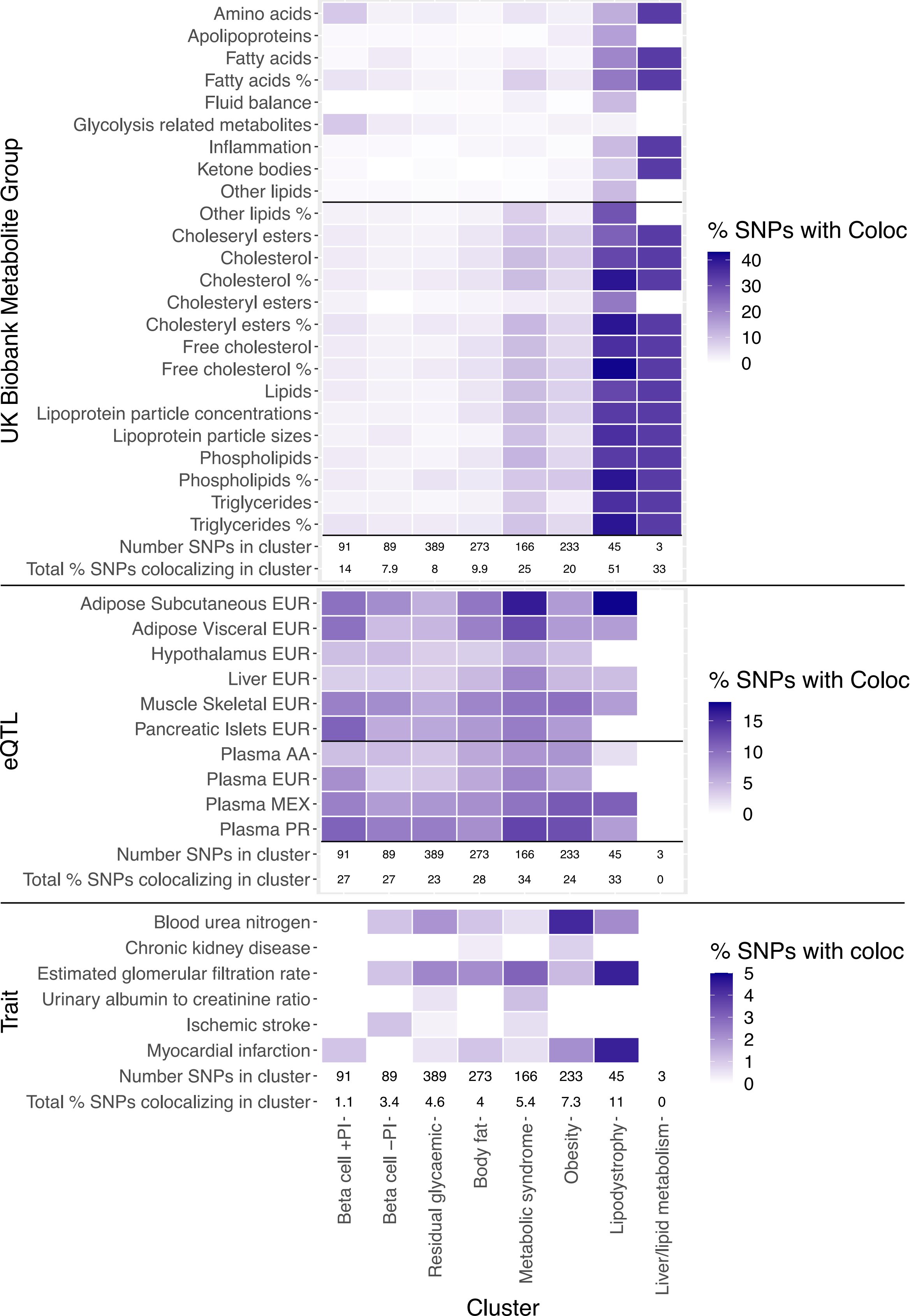

### Extended Data Figure 5

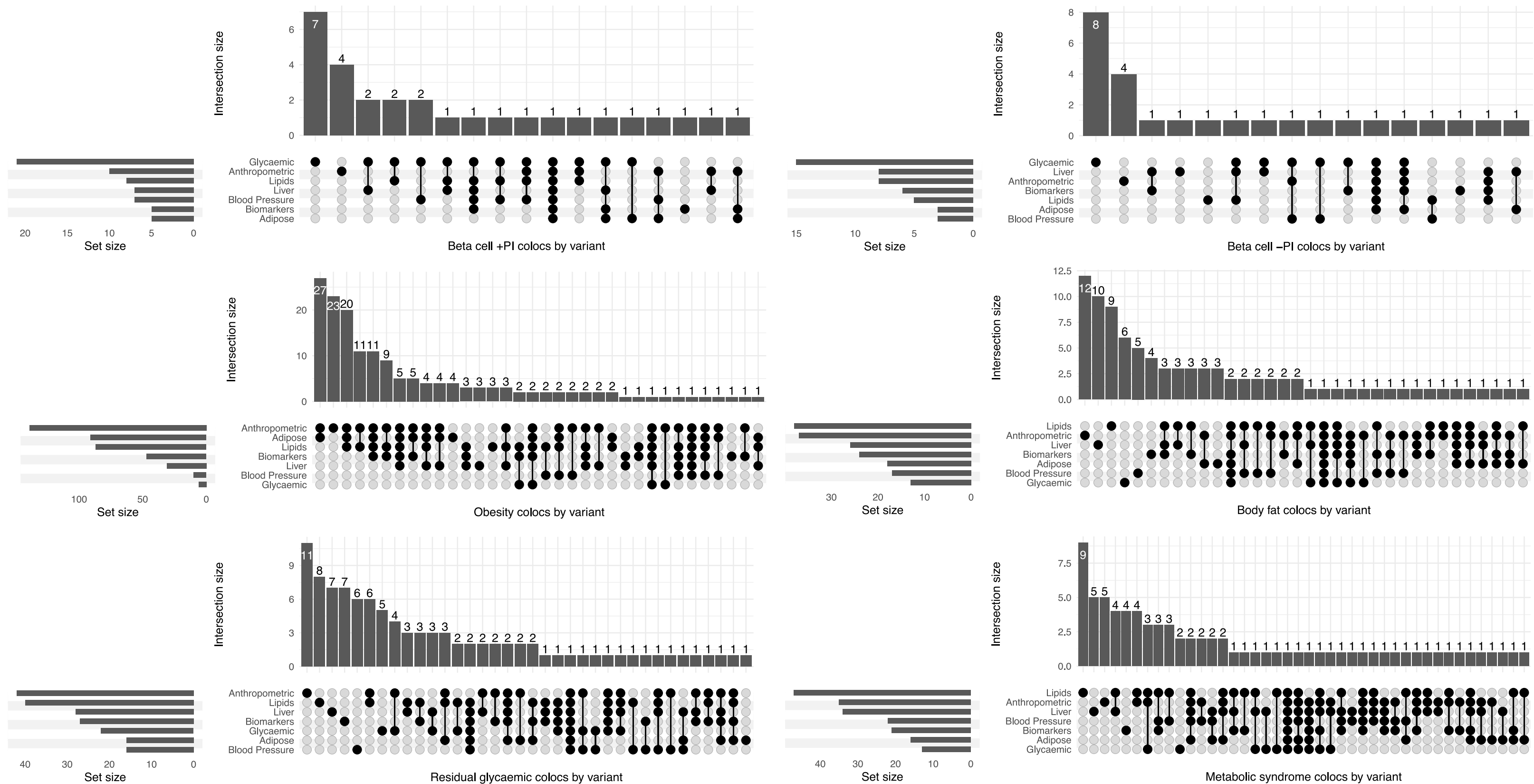

### Extended Data Figure 6

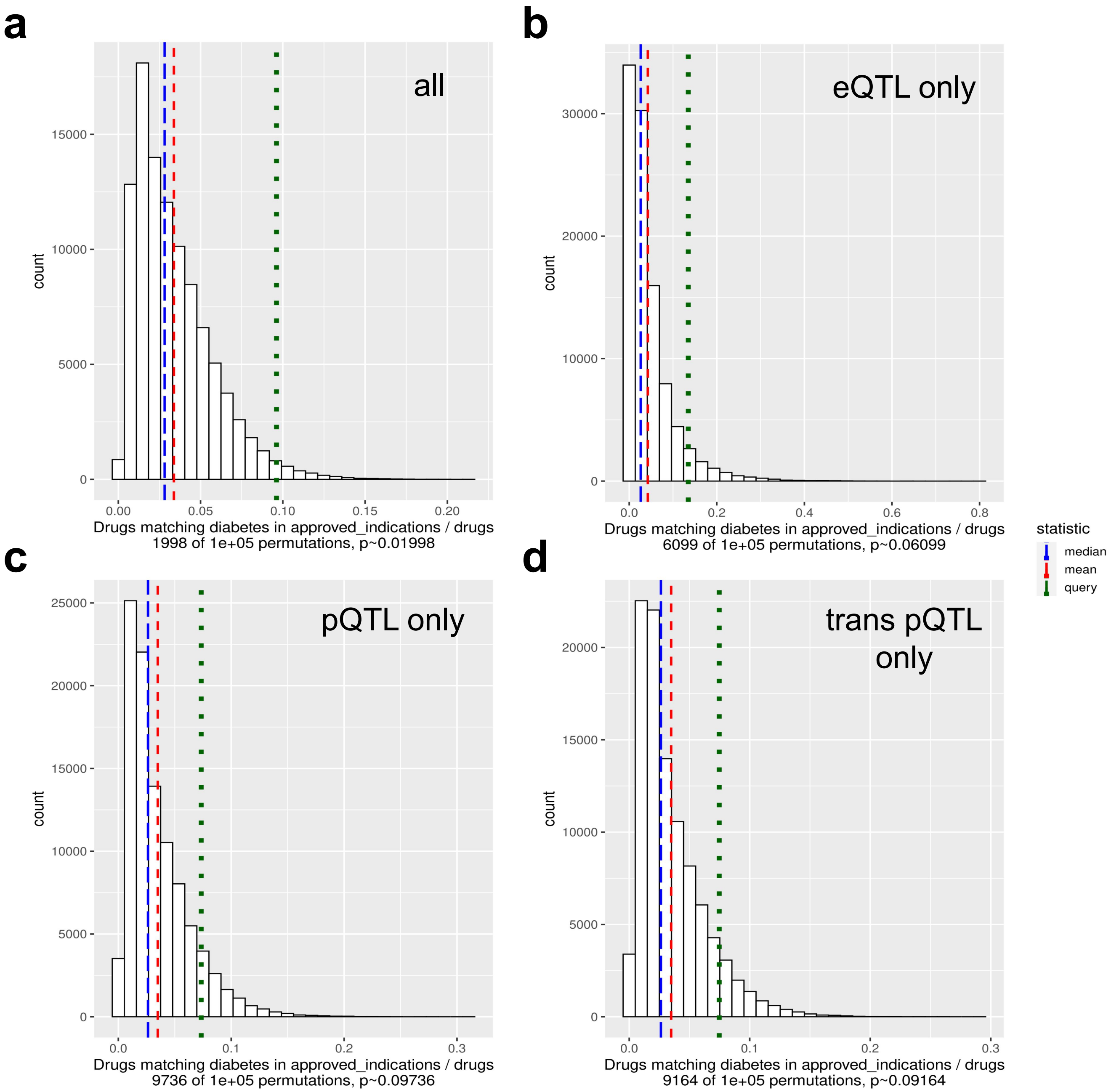
