## Extended Data Figure 7 for "Multi-omics characterization of type 2 diabetes associated genetic variation"

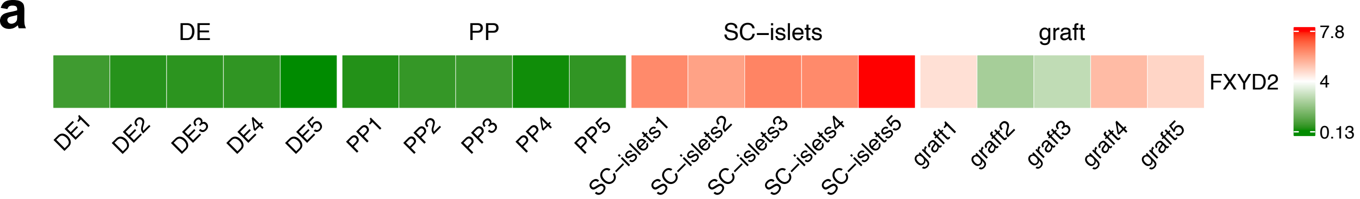


Gene counts were normalized as transcript per million (TPM)

| Gene | DE1 | DE2 | DE3 | DE4 | DE5 | PP1 | PP2 | PP3 | PP4 | PP5 | SC-islets1 | SC-islets2 | SC-islets3 | SC-islets4 | SC-islets5 | graft1 | graft2 | graft3 | graft4 | graft5 |
|---|---|---|---|---|---|---|---|---|---|---|---|---|---|---|---|---|---|---|---|---|
| FXYD2 | 0.55 | 0.28 | 0.33 | 0.37 | 0.09 | 0.25 | 0.42 | 0.50 | 0.14 | 0.38 | 74.76 | 54.95 | 80.78 | 74.78 | 221.89 | 23.05 | 4.08 | 6.00 | 38.79 | 27.12 |

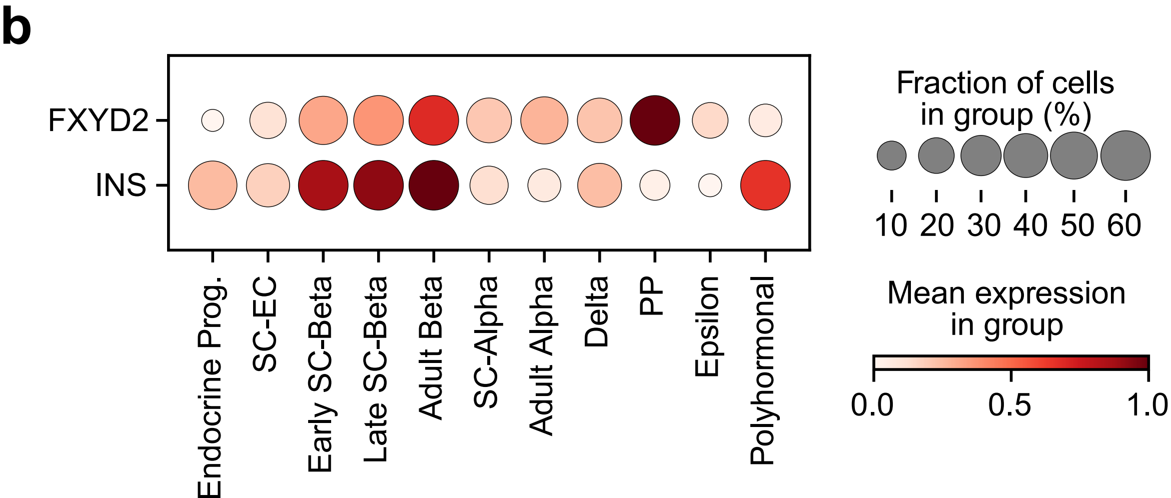
